# Patient Perception of Endoscopic and Medical Therapies for Weight Loss

**DOI:** 10.1101/2023.09.26.23296199

**Authors:** Monica Saumoy, Yinglin Gao, Kelly Allison, Peter F. Cronholm, Octavia Pickett-Blakely, Michael L. Kochman, Nikhil R. Thiruvengadam

## Abstract

**Background:** For weight management, patient choice has increasingly driven therapeutic options, with less than 1% of eligible patients choosing bariatric surgery. The aim of this survey was to understand patient perceptions of endoscopic bariatric therapies (EBTs) and obesity-based pharmacotherapy.

**Methods:** An anonymously collected 7-question survey was distributed to gastroenterology patients undergoing screening colonoscopy.

**Results:** A total of 184 patients participated in the survey. Participants demonstrated a greater lack of knowledge of EBTs (78.2% unaware) compared to pharmacotherapy (35.9% unaware). 40.8% of respondents perceived that EBTs were not available in the United States. Only 15.8% of respondents recognized that pharmacotherapy requires long-term treatment to maintain weight loss. Disparities were noted in primarily Spanish-speaking patients demonstrating a lower awareness of the availability of EBTs in the US (57.6% unaware) and pharmacotherapy (62.1% unaware) compared to those with English as the primary language.

**Conclusions:** Patient knowledge and preference are key to engaging in weight loss therapies. Knowledge gaps regarding weight-loss options, particularly EBTs, can limit the utilization of all options for the care of patients with obesity.

---

Rates of obesity have risen exponentially, both in the US and worldwide. In the US, current models predict that by the year 2030, nearly half of adults will have obesity, and nearly a quarter of adults will have severe obesity.^1^ Current weight loss tools include lifestyle intervention, pharmacotherapy, bariatric surgery, and endoscopic bariatric therapies (EBTs). Compared to lifestyle intervention, bariatric surgery has been shown to achieve superior weight loss outcomes, improvement in metabolic comorbidities, and reduced mortality.^2^ But, despite the evidence that bariatric surgery is effective, less than 1% of eligible patients choose to undergo bariatric surgery in the US.^3^

Pharmacotherapy provides an alternative weight management strategy, with a variety of medications demonstrating successful weight loss. Most recently, semaglutide, a glucagon-like peptide-1 agonist, has shown the most patient-driven interest with an average of 6-17% total body weight loss (TBWL) along with improvement in markers of cardiovascular health.^4^ Another agent, tirzepatide, has demonstrated an average of 15-20.5% TBWL.^5^ Unfortunately, access to pharmacotherapy has been limited due to significant demand, supply chain issues, and lack of insurance coverage outside of the diagnosis of diabetes.

Nationally, there has also been increasing utilization of EBTs, though not as significant as pharmacotherapy. EBT procedures such as endoscopic sleeve gastroplasty (ESG) and intragastric balloon, have previously been limited by the lack of insurance coverage. But the recent C code designated by the Centers for Medicare and Medicaid (CMS) for ESG after the MERIT trial^6^ demonstrated clinical success, and has allowed for increased access. Though there has been a significant body of research demonstrating the effectiveness of EBTs, there has not been much investigation on improving patient engagement and implementation, as compared with bariatric surgery.^7, 8^

Patients will often turn to their primary care physician (PCP) for advice on how to proceed with weight management. PCPs are increasingly being asked by patients to prescribe pharmacotherapy for weight loss. And a recent survey study demonstrated that 76.2% of PCPs discussed bariatric surgery with patients, but 52.3% of PCPs were not aware of EBTs as weight loss interventions.^9^ A critical issue is whether patients’ knowledge of EBTs is similarly restricted, limiting options in shared decision-making discussions with PCPs and engagement with gastroenterologists. This survey aimed to determine patients’ knowledge of EBTs and pharmacotherapy.

We performed a questionnaire-based, cross-sectional study of patients at Penn Medicine Princeton Health and at Loma Linda University system in August 2023. Institution of research ethics board (IRB) approval was obtained. The questionnaire was developed to determine patient knowledge of endoscopic bariatric therapies and pharmacotherapy for weight loss. Items were generated following a review of the literature and study team discussion. Surveys were administered in English and Spanish to patients presenting for outpatient endoscopy – as a survey of non-targeted patients of the general population. Survey responses were anonymous and managed using REDCap electronic data capture tools. No identifiable data were collected for the survey respondents and no incentives were provided for the survey to participants. After completion of data collection, quantitative analysis was performed using Stata software version (Stata Corp., 2017, Stata statistical software: Release 15. College Station, Texas).

184/229 (80.3%) patients completed the survey, which was collected over a one-month period. Spanish was the primary language for **36%** of respondents. (Table 1) Most of the respondents (**78**.**3%**) were unaware of endoscopic procedures to help patients lose weight. A fraction of respondents had heard of the ESG (26.1%) or intragastric balloon (29.9%). A total of 40.7% of respondents believed that EBT was not available in the US. In contrast, 64.1% of respondents had previously heard of medications for weight loss. Though only 15.8% recognized that medications for weight loss needed to be continued long-term for maintenance. And only 34.2% of respondents stated that pharmacotherapy could successfully improve obesity-related medical comorbidities. Spanish-speaking individuals had lower rates of awareness of ESG (89% unaware), intragastric balloon (84.8% unaware), or knowledge of effective pharmacotherapy (62.1% unaware) compared to English-Speaking individuals. (Figure 1)

**Figure 1.**
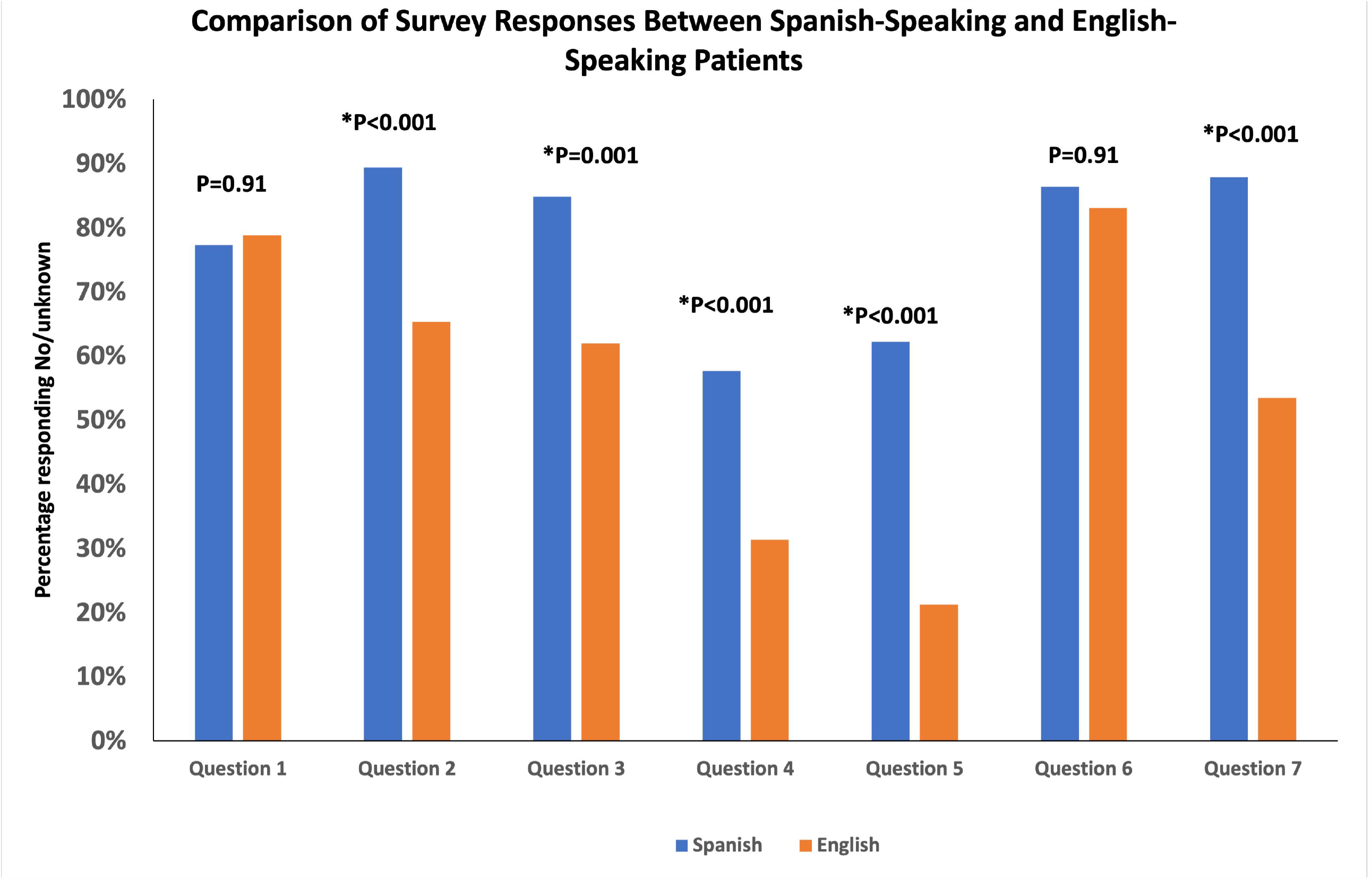
Bar graph demonstrating patient responses to survey questions separated between English and Spanish-speaking patients.

This survey investigated patient knowledge regarding endoscopic therapies for weight management. The survey demonstrated that a majority of patients (78.2%) were unaware of EBTs for weight management and had not heard of either ESG or intragastric balloon. Though a significantly higher proportion of patients (64.1%) were aware of medication therapy for weight loss, most respondents were not aware that weight-loss medications require long-term use for weight-loss maintenance.

Management of obesity and related complications has been an increasingly common indication for medical visits due to the obesity epidemic. Providers recognize that obesity is a complex disease that requires multimodal therapy, including clinicians specializing in medical, surgical, and endoscopic therapies, as well as dieticians, exercise therapists, and behavioral psychologists. However, access to such comprehensive centers is limited due to provider availability. If multidisciplinary centers are unavailable, referral patterns are driven by providers to singular centers, such as a bariatric surgery group or a gastroenterologist for endoscopic bariatric therapy. Despite the reported efficacy of EBTs, there is still a limited number of patients being referred for gastroenterologist evaluation. Knowledge gaps and consumer demand driving PCP referral patterns are likely responsible, as a recent survey study demonstrated that while 72.6% of PCPs discussed bariatric surgery as an option for weight loss, 52.3% of PCPs were not aware of EBTs, and only 24.6% of PCPs were familiar with the indications for EBTs.^9^ PCPs are typically the first medical providers to initiate patient discussions about obesity and the indications for weight loss. Therefore, provider education on EBTs will be key for improving patient uptake in weight loss therapies.

Patient preference is a significant factor to drive patient engagement and treatment decision-making. Patient preference is typically a combination of what therapies patients have been exposed to, either from their providers, friends/family, social media, or the lay press. Shared decision-making with patients and their providers is a key component of patient engagement. Patients who are engaged in chronic illness management, have demonstrated improvement in mortality.^10^ If patients are unaware of potential weight loss therapies, this can limit optimal care of this patient population. In our survey, only 21.7% of respondents recognized that EBT is an option for weight loss, demonstrating a significant knowledge gap. The knowledge gap widens for patients for whom English is not their primary language. This survey adds to the current body of literature that there are significant disparities related to language barriers in healthcare, which limits compliance and successful clinical outcomes.^11, 12^ Patient engagement tools and decision aids have been utilized in other clinical scenarios to support and help patients make medical decisions and include key clinical content for patient understanding.^13^ This suggests that targeted educational programs, in their primary language, to educate patients on comprehensive treatment options for weight loss (surgery, pharmacotherapy, and EBTs) may improve patient access and engagement.

Pharmacotherapy provides an adjunctive weight management strategy, with a variety of medications demonstrating successful weight loss. In addition to success in clinical trials, there have been numerous reports in the media about celebrity use of pharmacotherapy, which has driven overall public interest. However, it is unclear what the public understands regarding pharmacologic weight management. Specifically, medication therapy needs to be continued indefinitely as demonstrated in the STEP4 trial which showed rapid weight regain when semaglutide was discontinued.^14^ In our study, only 15.8% of respondents recognized that pharmacotherapy requires long-term use to maintain weight loss. However, there are serious cost concerns with the use of these medications, resulting in limited insurance coverage in the US. A recent cost-effectiveness study demonstrated that long-term therapy with semaglutide is currently not cost-effective at a willingness-to-pay threshold of $100,000/QALY due to its high annual cost.^15^ Indeed, the National Institute for Health Care Excellence in the UK approved only a limited 2-year course of semaglutide based on an industry-sponsored analysis demonstrating that a 2-year course of semaglutide was cost-effective compared to lifestyle intervention in patients with obesity and at least one comorbidity.^16^ But as suggested by the STEP4 trial, planned discontinuation has the risk of rapid weight regain. The risk of rapid weight cycling is likely increased by multiple factors: poor insurance coverage, high out-of-pocket costs, limited access to long-term use of the medication, and lack of patient knowledge of the need to continue pharmacotherapy to maintain long-term weight loss. In turn, rapid weight cycling may increase depressive symptoms,^17^ risk of cardiovascular disease, and all-cause mortality.^18^

Due to the rise in obesity and the simultaneous need for weight loss therapies, engagement by all key stakeholders is necessary for the management of patients with obesity and obesity-related complications. However, knowledge gaps regarding EBTs, incorrect and inadequate perceptions of pharmacologic weight therapies, limited access to dedicated bariatric centers, and lack of financial coverage, can limit the optimal care of this patient population. Targeted educational programs to educate patients and providers on the most current guidelines and weight loss interventions may improve patient access and engagement. While knowledge and awareness are critical aspects of behaviors, much more is required to support adaptive changes in behaviors that can result in meaningful and sustained weight loss.

## Data Availability

All data produced in the present work are contained in the manuscript

## Abbreviations used in this paper

EBT: (endoscopic bariatric therapies)
ESG: (endoscopic sleeve gastroplasty)
TBWL: (total body weight loss)

## Captions

Table 1. Survey administered to patients with collected responses. Survey responses tabulated as all responses and responses of patients with Spanish as their primary language.

## References

1. Ward ZJ, Bleich SN, Cradock AL, et al. Projected U.S. State-Level Prevalence of Adult Obesity and Severe Obesity. N Engl J Med 2019;381:2440–2450.

2. Chang D-M, Lee W-J, Chen J-C, et al. Thirteen-year experience of laparoscopic sleeve gastrectomy: surgical risk, weight loss, and revision procedures. Obesity surgery 2018;28:2991–2997.

3. Martin M, Beekley A, Kjorstad R, et al. Socioeconomic disparities in eligibility and access to bariatric surgery: a national population-based analysis. Surg Obes Relat Dis 2010;6:8–15.

4. Gao X, Hua X, Wang X, et al. Efficacy and safety of semaglutide on weight loss in obese or overweight patients without diabetes: A systematic review and meta-analysis of randomized controlled trials. Front Pharmacol 2022;13:935823.

5. Jastreboff AM, Aronne LJ, Ahmad NN, et al. Tirzepatide Once Weekly for the Treatment of Obesity. N Engl J Med 2022;387:205–216.

6. Abu Dayyeh BK, Bazerbachi F, Vargas EJ, et al. Endoscopic sleeve gastroplasty for treatment of class 1 and 2 obesity (MERIT): a prospective, multicentre, randomised trial. Lancet 2022;400:441–451.

7. Chee A, Abdel-Rasoul M, Zoretich K, et al. Bariatric Patient Engagement in a Presurgery Virtual Patient Navigation Platform (VPNP). Obes Surg 2023.

8. Musbahi A, Clyde D, Small P, et al. A Systematic Review of Patient and Public Involvement (PPI) in Bariatric Research Trials: The Need for More Work. Obes Surg 2022;32:3740–3751.

9. Ouni A, Khosla AA, Gomez V. Perception of Bariatric Surgery and Endoscopic Bariatric Therapies Among Primary Care Physicians. Obes Surg 2022;32:3384–3389.

10. Krist AH, Tong ST, Aycock RA, et al. Engaging Patients in Decision-Making and Behavior Change to Promote Prevention. Stud Health Technol Inform 2017;240:284–302.

11. Rosen CB, Roberts SE, Sharpe J, et al. A study analyzing outcomes after bariatric surgery by primary language. Surg Endosc 2023;37:6504–6512.

12. Kroner Florit PT, Corral Hurtado JE, Wijarnpreecha K, et al. Bariatric Surgery, Clinical Outcomes, and Healthcare Burden in Hispanics in the USA. Obes Surg 2019;29:3646–3652.

13. Stacey D, Legare F, Lewis K, et al. Decision aids for people facing health treatment or screening decisions. Cochrane Database Syst Rev 2017;4:CD001431.

14. Rubino D, Abrahamsson N, Davies M, et al. Effect of continued weekly subcutaneous semaglutide vs placebo on weight loss maintenance in adults with overweight or obesity: the STEP 4 randomized clinical trial. Jama 2021;325:1414–1425.

15. Saumoy M, Gandhi D, Buller S, et al. Cost-effectiveness of endoscopic, surgical and pharmacological obesity therapies: a microsimulation and threshold analyses. Gut 2023.

16. National Institute for Health and Care Excellence. (2022). Semaglutide for managing overweight and obesity https://www.nice.org.uk/guidance/gid-ta10765/documents/final-appraisal-determination-document.

17. Quinn DM, Puhl RM, Reinka MA. Trying again (and again): Weight cycling and depressive symptoms in U.S. adults. PLoS One 2020;15:e0239004.

18. Oh TJ, Moon JH, Choi SH, et al. Body-weight fluctuation and incident diabetes mellitus, cardiovascular disease, and mortality: a 16-year prospective cohort study. The Journal of Clinical Endocrinology & Metabolism 2019;104:639–646.

