## Supplementary material for "Patient Perception of Endoscopic and Medical Therapies for Weight Loss": Table 1

| Question | All Responses  N=184 | | Response (Spanish Survey)  N=66 | |
| --- | --- | --- | --- | --- |
|  | Yes/true (%) | No/false or Unknown (%) | Yes/true (%) | No/false or Unknown (%) |
| There are endoscopic procedures to help patients lose weight loss. | 21.7 | 78.3 | 22.7 | 77.2 |
| Have you heard of the Endoscopic Sleeve Gastroplasty? | 26.1 | 73.9 | 10.6 | 89.3 |
| Have you heard of the Intragastric Balloon? | 29.9 | 70.1 | 15.2 | 84.8 |
| Endoscopic methods of weight loss are available in the United States. | 59.2 | 40.8 | 42.4 | 57.6 |
| Have you heard of medications for weight loss? (for example Ozempic or Wegovy) | 64.1 | 35.9 | 37.9 | 62.1 |
| If you start medications for weight loss, they need to be continued indefinitely for weight loss maintenance. | 15.8 | 84.2 | 13.6 | 86.4 |
| Medications for weight loss do not improve other conditions such as diabetes or hypertension | 34.2 | 65.8 | 12.1 | 87.9 |
